# Using wastewater-based epidemiology with local indicators of opioid and illicit drug use to overcome data gaps in Montana

**DOI:** 10.1101/2020.04.18.20064113

**Authors:** Miranda Margetts, Aparna Keshaviah, Xindi C. Hu, Victoria Troeger, Jordan Sykes, Nicholas Bishop, Tammy Jones-Lepp, Marisa Henry, Deborah E. Keil

## Abstract

Opioid misuse takes an enormous toll on the health and economy of the U.S., costing 130 lives per day, and an estimated 157 billion dollars per year. Mitigating the epidemic requires better information on the mix of drugs being used in a community and when and where drug use concentrates. Many widely used data sources, such as national population surveys, have a two-year lag before the data are available, and underestimate the prevalence of drug use because they rely on self-reported information about a stigmatized behavior. For rural populations, the data gap is compounded by privacy policies that suppress key information (for example, opioid-related death rates) in small populations. Municipal wastewater testing is an innovative approach to assessing community-level drug use that can provide near real-time, cost-effective, and unbiased measures of drug use. Importantly, wastewater-based data cannot reveal who is using a particular drug, mitigating privacy concerns. The methodology is routinely used across Europe and Australia as part of a holistic multi-indicator monitoring and alert system. When combined with geospatial mapping and advanced analytics, wastewater-based data can help locate geographic hotspots of use and provide an early warning for new substances entering into a community. We conducted a pilot study to compare temporal trends in estimates of per-capita drug use between wastewater-based data and other local data sources in two municipalities (one urban, one rural) in Montana. In the first phase of our study (Bishop et al, 2020, *manuscript is in submission to Sci Total Environ*), we optimized testing to estimate per-capita drug use in wastewater in an urban and rural municipality. Here, we report the value of these data for public health officials to predict rather than react to changes in drug use patterns. We also illustrate how rural populations can use the methodology to assess the impacts of local law enforcement efforts and evaluate the effectiveness of programs, such as the prescription take-back program, to improve public health and safety.

## Introduction

Governments and health care providers across the U.S. continue to search for a holy-grail community monitoring mechanism to gauge substance use trends in real time. Given the urgency and economic burden associated with the opioid epidemic, recently estimated at $157B per year (Davenport et al, 2019), it is critical to identify a better data source—one that can provide more timely, objective, and comprehensive measures of drug use than currently exist. Many widely used data sources, such as national population surveys, have a two-year lag before the data are available, and underestimate drug use prevalence because they rely on self-reported information about a stigmatized behavior. For rural populations, the data gap is compounded by privacy policies that suppress health measures (for example, opioid-related death rates) in small populations and, despite higher rates of overdose among rural opioid users, rural users are less likely to report recent risk behaviors (Dunn et al, 2016). Montanan health care providers wrote 90 opioid prescriptions per 100 persons in 2015 (approximately 722,011 prescriptions) which exceeds the average national rate of 70 opioid prescriptions per 100 persons (NIDA, 2018). Limited data sharing between public health, primary care, hospital systems, first responder units and law enforcement agencies further hampers the ability to obtain an accurate picture of illicit drug use and prescription diversion.

Advances in wastewater testing and wastewater-based epidemiology are being utilized in response to this unmet demand (Castiglioni et al, 2014; Daughton, 2001; Banta Green and Field, 2011; Zuccato et al, 2008). Municipal wastewater is a combination of household sewage, industrial run-off, and (in some locations) storm water. About 15,000 wastewater treatment plants (WWTPs) around the country collect wastewater from 81% of U.S. households (Department of Homeland Security, 2015). Wastewater testing (typically at the central WWTP) involves a three-stage process: (1) routine wastewater sampling, (2) chemical analysis (via mass spectrometry) to quantify the concentration of a drug (parent compound or its metabolites) in each sample, and (3) back-calculations to convert drug concentrations in each sample to a per-capita estimates of drug use in the WWTP catchment population (Castiglioni and Vandam, 2016). Over the past decade, municipal wastewater testing has been validated as an effective tool for tracking and identifying trends in prescription and illicit drug use (Castiglioni et al, 2013, van Nuijs et al, 2011), and one that sidesteps privacy concerns typically associated with behavioral health data (Hall et al, 2012).

Various studies report the successful detection of prescription and illicit drugs in community wastewater and illustrate how, when wastewater data are combined with data from syringe distribution programs, pharmacy prescriptions, and population surveys, data triangulation can be used to pinpoint illicit opioid use (Been et al, 2015, van Nuijs et al, 2015). However, to date, little research has been conducted in the U.S. to determine the feasibility of acquiring such complementary local data—particularly in rural areas, where data access can be challenging— and few studies have reported how wastewater data can yield insights for policymakers and public health officials.

Wastewater data can provide important information to characterize the spatial and temporal variations of community drug use, particularly when considered alongside local data. Here we describe a 10-week pilot study we developed in Montana. We examined the alignment between wastewater data and local indicators of drug use in two Montana municipalities (one urban, one rural) using data on emergency medical services (EMS) overdose calls, law enforcement drug seizures, and pharmacy prescriptions filled. Based on our findings, we describe how wastewater testing can help communities predict rather than react to changes in drug use, and illustrate how rural communities can use the methodology to assess the impact of local law enforcement efforts to improve public safety. Given recent federal funding investments in cross-sector data sharing and wastewater-based epidemiology, our study provides a timely validation of this innovative approach for detecting new drug threats and changes in community drug use over time.

## Methods

### Wastewater-based estimates of drug doses consumed

Wastewater-based estimates of drug use came from a wastewater pilot study undertaken in Montana (Bishop et al, 2020, *manuscript is in submission to Sci Total Environ*). The study involved collecting weekly 24-hour composite samples of untreated wastewater over a 10-week period (April 15 to June 20, 2019) at two municipal WWTP sites (Site A, Site B). Site A has a catchment population of 45,250 and is in an urban city, while Site B has a catchment population of 4,000 and is in a rural town. Solid phase extraction and liquid chromatography with dual mass spectrometry (LCMSMS) analysis was undertaken on each sample at Assurity Labs in Las Vegas, NV. Based on the LCMSMS analyses, the following 18 compounds were consistently detected at reliable concentrations across Site A and Site B: amphetamine, methamphetamine, 3,4-methylenedioxyamphetamine (MDA), 3,4-methylenedioxymethamphetamine (MDMA), morphine, 6-acetylmorphine, methadone, *2-Ethylidene-1,5-dimethyl-3,3-diphenylpyrrolidine* (EDDP), codeine, benzoylecgonine, hydrocodone, hydromorphone, oxycodone, noroxycodone, ketamine, fluoxetine, tramadol, and ritalinic acid. The measured drug concentrations were then subject to a series of back-calculations to determine the dose estimate in mg/person per day from the measured concentrations. (*A detailed methodology of the analytical techniques and calculations undertaken on the measured concentrations will be forthcoming in Bishop et al, 2020*).

### Local indicators of drug use

We obtained local data on drug use indicators from emergency medical services (EMS) call logs, prescription dispensing data, and drug seizure logs. Table 1 illustrates the data source, unit of measurement, availability of data across the site and the population coverage per the unit of measurement. We limited our analysis of these data sources to events that occurred during the wastewater sampling period, and which involved drugs that were included in the wastewater pilot study testing panel. Sourcing the local data involved collaborative efforts with community partners and state agencies involved in health care as it pertained to drug use, including the Montana Department of Public Health and Human Services, the Montana Statistical Analysis Center, community healthcare clinic and pharmacy collaborators.

**Table 1:** Local data sources used as community drug use indicators.

| Data source | Unit of measurement | Data availability | Population coverage per unit |
| --- | --- | --- | --- |
| Wastewater treatment plant | Wastewater sample | Sites A and B | Site A: 45,250 (42% of county)<br>Site B: 4,000 (33% of county) |
| Pharmacy | Filled prescription | Site B | 1 person |
| Law enforcement | Drug seizure | Site A | 1 – 250 people |
| Emergency medical services | Drug overdose call | Sites A and B | 1 person |

### Pharmacy prescriptions filled

As data from the Montana Prescription Drug Monitoring Program were not available at the time of our analysis, we instead obtained weekly logs of prescriptions filled at one of two pharmacies operating within the county where Site B is located. The logs included drug names/formulations, counts, and quantities of prescriptions filled for each week during the study period. The prescription drug names were mapped to their metabolites to enable cross-referencing with the wastewater-based data. To facilitate comparisons, we summarized, for each wastewater sample, the prescriptions filled during the week just prior to the wastewater sample collection date. We analyzed prescriptions filled before the wastewater collection date (rather than after) because sample collection at the WWTP reflects drug use occurring in the recent past (factoring in the time it takes for wastewater to flow through collection pipes and into the plant).

### Law enforcement drug seizures

Drug seizure logs were available for Site A only, and included information on the seizure date, drug type, quantity seized, and units corresponding to that quantity (which we assumed to be grams, if not specified). Our analysis focused on amphetamine/methamphetamine seizures (a combined category used in the logs); for all other drugs seized, there were either very few seizures during the study period, very few wastewater measurements above the limit of detection (LOD), or no testing for that substance in wastewater. We also focused on the top quartile of seizures based on the grams of drugs seized, as we hypothesized that large seizures might take enough drugs out of circulation to affect population-level estimates that wastewater data provide. We superimposed information on when these seizures occurred on top of wastewater trend lines for amphetamines and methamphetamines.

### EMS drug overdose calls

For the counties in which Site A and Site B are located, we obtained de-identified logs of calls placed to EMS for drug overdoses. The call logs included the call date, county, whether or not the overdose occurred in the collection region corresponding to the WWTP service area, and the list of drugs involved (which we coded into indicators for each drug of interest).

## Results

Figure 1 shows how the community drug use profiles differed by site, with color-coding used to indicate the drug class (opioid, stimulant or anti-depressant). The three most commonly-used drugs were the same across both sites: tramadol, methamphetamine and codeine. However, methamphetamine use was six to seven times higher in Site B than Site A, and three to five times higher than use of any opioids at either site. The heroin metabolite (6-acetylmorphine) was below the LOD for most samples at both sites, and MDA, MDMA, and ketamine concentrations were below the LOD for several samples at site B only. Trend lines of the estimated dose/person at each site revealed substantial fluctuation in drug use over the study period for most drugs. However, a few consistent patterns emerged: at both sites, there was a steady increase in hydrocodone use, which more than doubled over the study period; a spike in morphine, fluoxetine, and MDA use on May 22, 2019; and a spike in hydromorphone, noroxycodone, and ketamine use on June 5, 2019. After examining local events that might explain the upticks in use, we learned that June 5 corresponded with the week of a local high school graduation.

**Figure 1:**
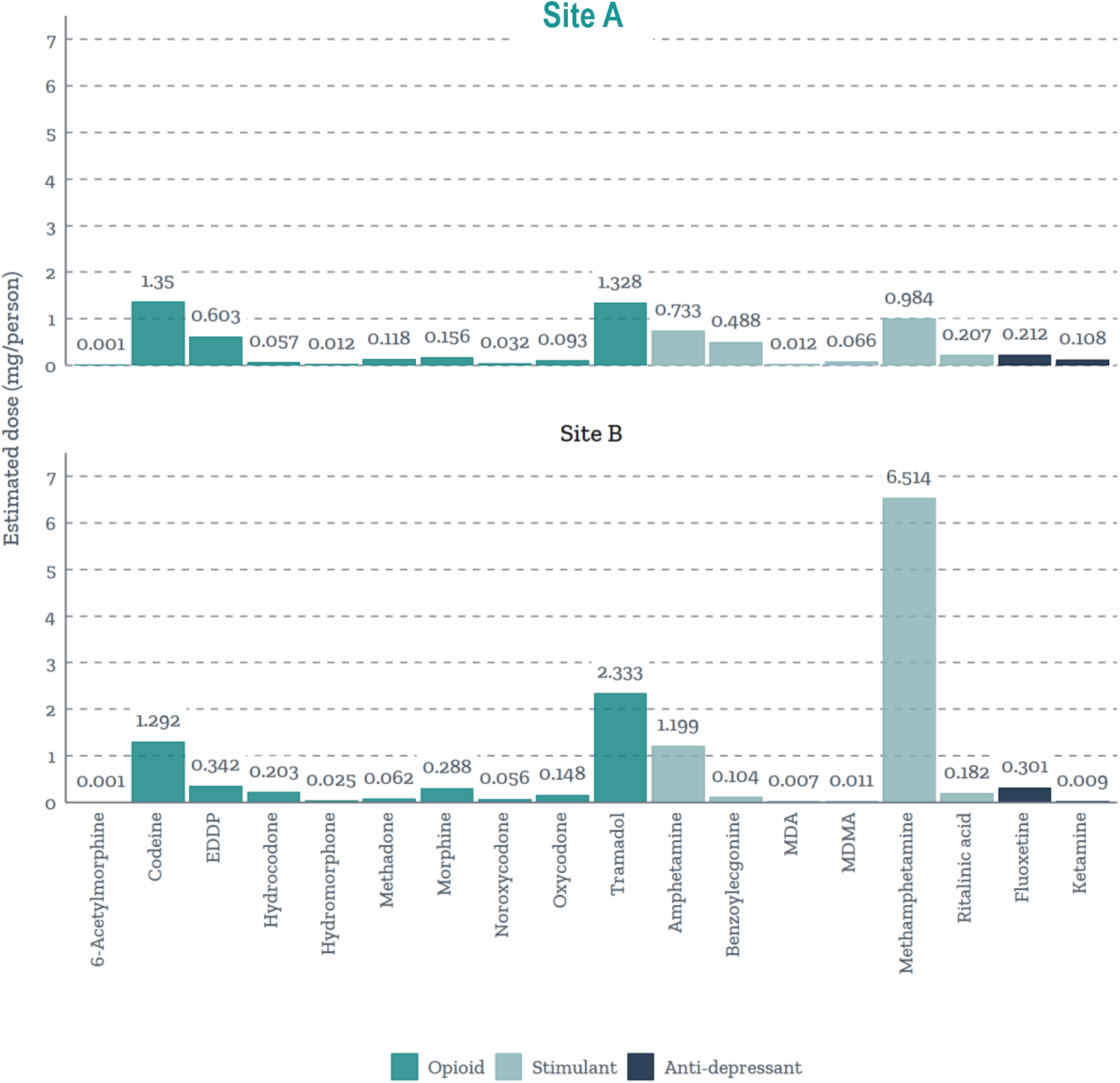
Wastewater-based estimates of the average dose (mg/person per/day, normalized to population size) per drug by site. The bar charts show differences in the magnitude and types of drugs used across Site A and Site B. The height of each bar represents the average estimated dose in mg/person p/day, normalized to population size, across the study period (4/15/2019 to 6/20/2019).

### Prescription data compared to wastewater drug estimates (Site B)

Trends in the number of drug doses in filled prescriptions aligned well with trends in number of drug doses excreted into the wastewater estimates for some drugs, though not all. Specifically, rises and dips in the use of amphetamines, codeine, EDDP (with the exception of the June 5 assessment), methadone (early in the study period), and morphine (with the exception of June 5) were fairly consistent between the wastewater and prescription data. Figure 2A shows the alignment in codeine use estimates. By contrast, Figure 2B suggests that methamphetamine use in the community may be much higher than expected based on pharmacy logs, perhaps indicating the magnitude of black-market methamphetamine use.

**Figure 2:**
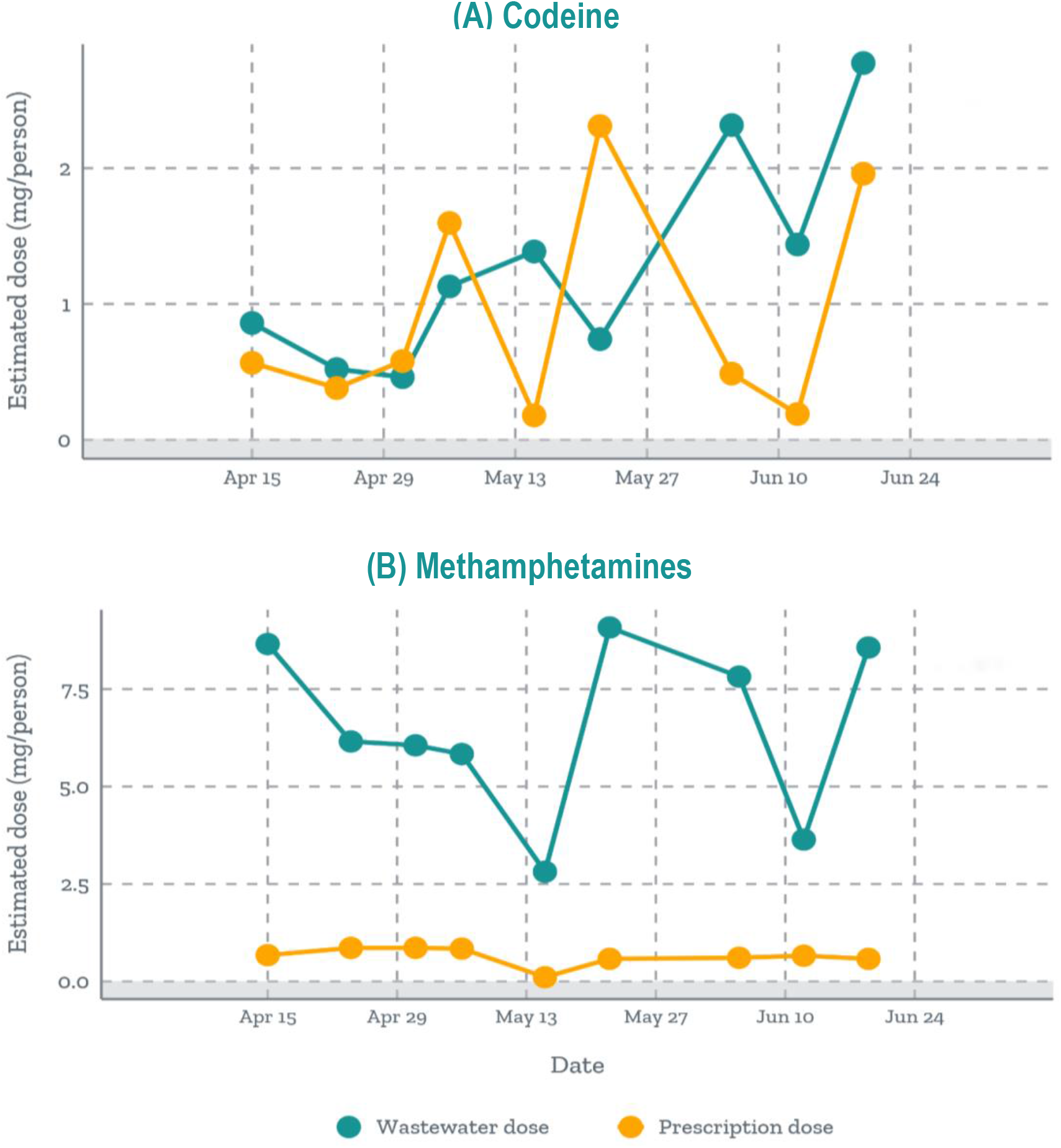
Wastewater vs. prescription data on trends in codeine and methamphetamine use. This graph overlays the number of doses of prescription (A) codeine and (B) methamphetamines filled at a local pharmacy versus the number of doses excreted into the wastewater at Site B. For each wastewater sample, we analyzed prescriptions filled during the week just prior to the wastewater sample collection date.

### Drug seizure data from law enforcement (Site A only)

Comparing wastewater data with drug seizure data revealed the impact of law enforcement activities on community drug use (Figure 3). After each major seizure of amphetamines/methamphetamines, the concentration of methamphetamines in wastewater declined, with two exceptions. Immediately after seizures that occurred on May 18 and June 24, the concentration of methamphetamines in wastewater rose. However, the concentration amphetamines in wastewater declined, thus maintaining the expected pattern.

**Figure 3:**
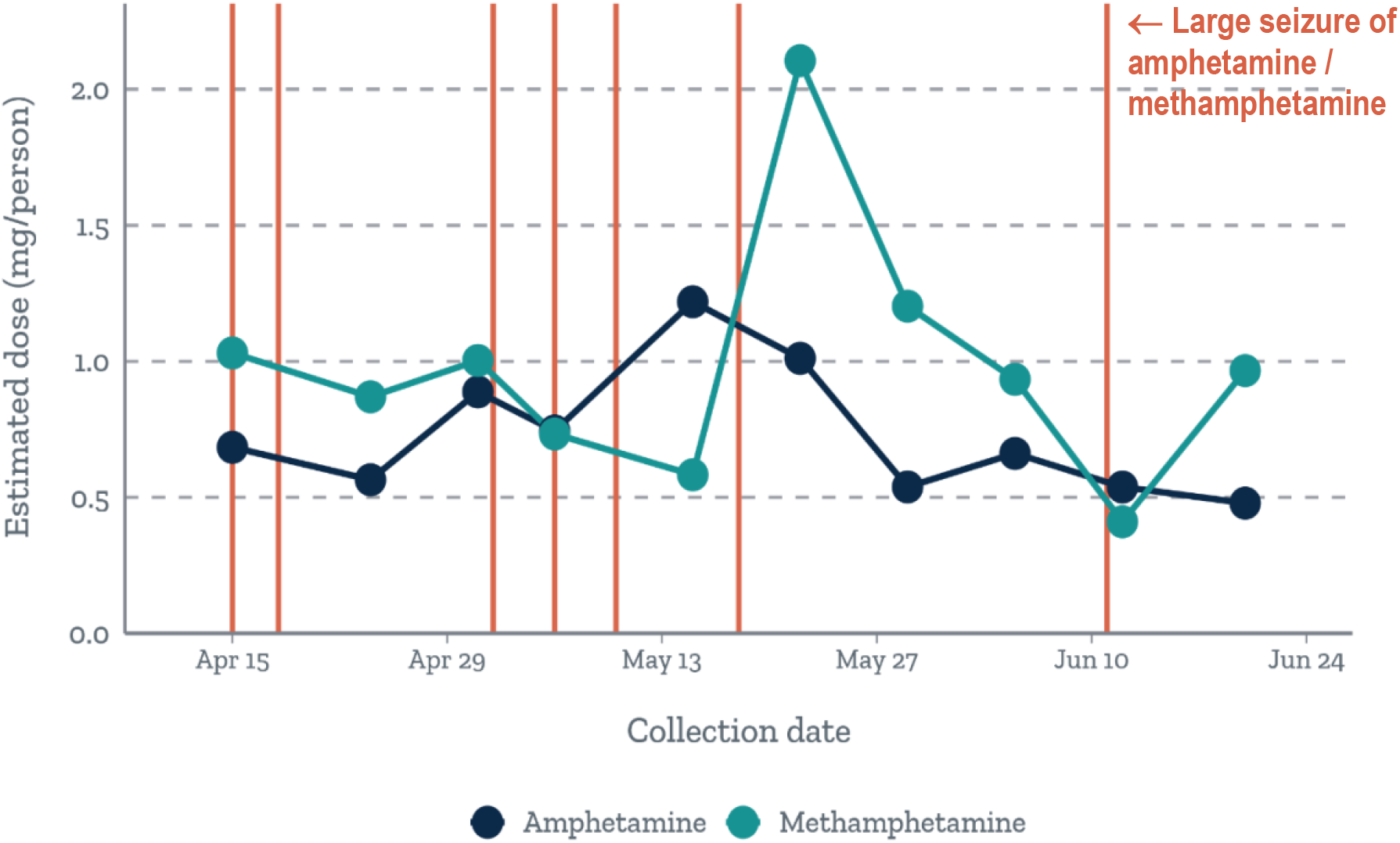
Using wastewater data to gauge the impact of drug seizures on community use of methamphetamines and amphetamines. Temporal trends in wastewater doses for Methamphetamines and Amphetamines; vertical lines indicate when large (top quartile) drug seizures of Amphetamines/Methamphetamines occurred during the study period.

### EMS call-logs (Site A and Site B)

The timing of EMS drug overdose calls in relation to wastewater concentrations of drugs suggest that wastewater data might be used to predict when a call to EMS for an overdose involving a particular drug might be expected. At both sites and for most drugs, EMS calls for an overdose involving a given drug occurred after community use of that drug was high, based on wastewater concentrations. Of the 12 calls (8 in Site A, 4 in Site B) placed during the study period that involved at least one targeted drug of interest, 7 occurred within 2 weeks after at least one of the drugs involved in the overdose had high or peak use in the community (i.e., wastewater doses that were 58% to 551% greater than the minimum dose measured during the study period), and 4 occurred within 2 weeks after moderate community use (i.e., wastewater doses that were 26% to 65% greater than the minimum dose measured during the study period). Figure 4 illustrates these trends for EMS calls that involved (A) tramadol and (B) heroin. Even though most of the wastewater samples we collected had heroin levels that were below the LOD, we found soon after community use spiked (resulting in a measurement above the LOD), an EMS call was placed for an overdose involving heroin.

**Figure 4:**
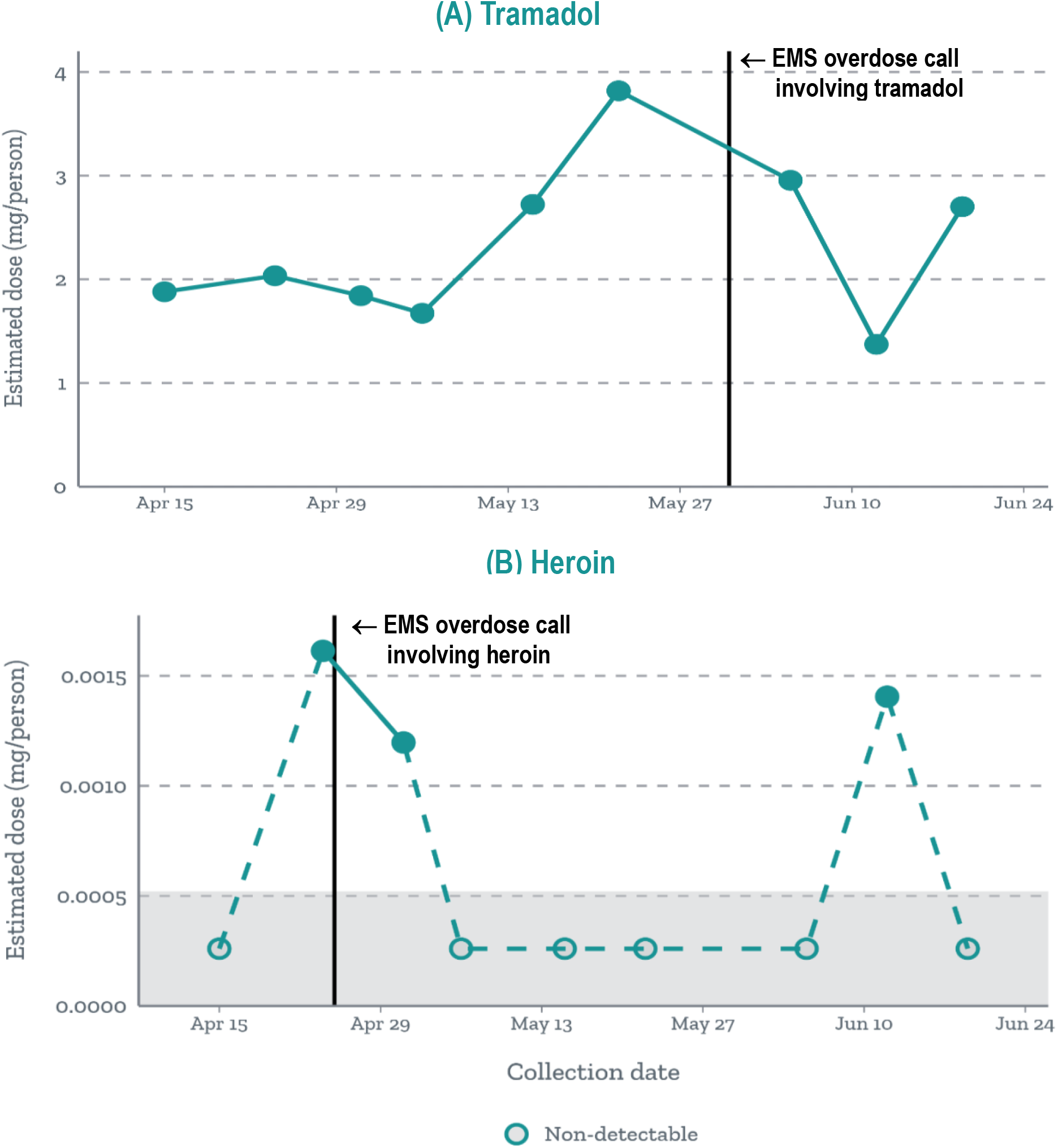
Assessing the timing of EMS overdose calls in relation to changes in wastewater estimates of tramadol and heroin use. Trends in wastewater dose estimates for (A) tramadol and (B) heroin metabolite (6-MAM) use at Site B are plotted over time, with vertical lines indicating when EMS calls for a drug overdose involving those substances occurred.

## Discussion

Wastewater-based profiles of drug use were similar across the urban and rural Montana communities studied. Our findings demonstrate the capacity of wastewater testing to capture community drug use trends, and align with trends in the literature. The high rates of tramadol use may reflect the noted surge in tramadol prescribing in the U.S. in recent years due to the perception that tramadol is less addictive than short-acting opioids (Thiels et al, 2019; Jeffrey et al, 2018). And the prevalence of methamphetamine use across both sites, and particularly in the rural community, aligns with a report from the Montana Department of Justice (2017) indicating a significant resurgence of methamphetamine use across the state in recent years. Although the opioid epidemic continues to be a major focus of federal funding and programs continue to target opioid use disorder (such as Medication-Assisted Treatment), wastewater data suggest the need to monitor on a broader array of drugs circulating in our communities, and to boost support for methamphetamine addiction. Moreover, our data suggest that that pharmacy prescription logs paint an incomplete picture of methamphetamine use, and that the illicit market may be responsible for recent surges around the country.

Our results also support the recently-described value of drug seizure data to complement wastewater-based data. Bruno et al (2018) illustrated how increases in the purity of methamphetamine seized within a wastewater catchment area may account for proportional increases in methamphetamine concentration loads reflected in wastewater samples. And the Australian Criminal Intelligence Commission has begun routinely using wastewater-based epidemiology to assess operational priorities (Australian Criminal Intelligence Commission, 2019). The drug seizure data recorded by law enforcement agencies within our investigation combined both amphetamine and methamphetamine in the same category. But by comparing drug seizure data with wastewater-based estimates of amphetamine and methamphetamine use, we could discern which seizures likely involved methamphetamines versus amphetamines, thus illustrating the value of wastewater data to provide detailed information on the effectiveness of law enforcement activities.

Various aspects of wastewater-based epidemiology require further optimization, given the novel nature of these investigations. Uncertainties exist in relation to the stability of the metabolites, back-calculations to estimate drug-dose consumed, population served and the excretion percentage (Castiglioni et al, 2013). For the wastewater data, identifying more specific or stable metabolites of some drugs (such as heroin) would improve detection (McCall et al, 2016). In the EMS overdose call and drug seizure logs, the types of drug involved in the EMS call and seizure logs are not consistently verified by a toxicological laboratory. The prescriptions filled do not necessarily represent prescriptions taken by individuals within the WWTP service areas. There is also uncertainty around the estimation of the population served, however studies are identifying population biomarkers, such as cotinine, that may improve this estimate (Cheng et al, 2014). Finally, the difficulty we encountered in acquiring a consistent set of local drug use indicators across both counties limited our ability to conduct more thorough comparisons, and to examine the generalizability of our findings.

## Conclusion

Healthcare professionals utilize a variety of data and tools for tailored, personalized medicine that aims to improve the health outcomes of their patients. Local public health data, such as air quality and disease outbreaks, and other health-related statistics routinely inform healthcare decision making at both the individual and community-level. However, the well-recognized limitations in data on prescription misuse and illicit drug use support the need for a better monitoring tool for communities.

Although still in its infancy in the U.S., there is rapidly growing interest in the insights that wastewater-based epidemiology data can provide. The methodology can be used to identify new drug threats entering into a community, inform how to tailor addiction treatment and prescribing practices according to shifts in drug use, evaluate the effectiveness of new programs or policies to reduce substance use, and provide a data-driven strategy for targeting limited resources. Going forward, for successful implementation of wastewater-based epidemiology, interagency collaborations that involve representatives from public health, medicine, behavioral health, law enforcement, and wastewater municipalities will be required (Keshaviah, 2017). Governance frameworks and best practices for utilization and implementation have yet to be established within the US, but because wastewater testing relies on a fairly standardized infrastructure for sampling and testing that is already in operation across the U.S., it is cost-efficient and scalable. Above all, because wastewater testing can provide unbiased, privacy-preserving measures of community drug use, it can empower healthcare professionals and public health officials to address the breadth and complexity of substance abuse in near real-time and help prevent the next drug epidemic.

## Data Availability

All data referred to is available at request to authors.

## Acknowledgements

Funding support was received from the National Institute of General Medical Sciences of the National Institutes of Health *(Award Number U54GM115371. PI: Dr. Deborah Keil)*.

The Institutional Review Board (IRB) of Montana State University provided IRB exemption for this research.

## References Cited

Australian Criminal Intelligence Commission. (2019). National Wastewater Drug Monitoring Program—Eighth Report. Retrieved from https://www.acic.gov.au/publications/reports/national-wastewater-drug-monitoring-program-eighth-report

Banta-Green, C., & Field J. (2011). City-wide drug testing using municipal wastewater: A new tool for drug epidemiology. Significance: The Royal Statistical Society. June, 2011.

Béen, F. (2015). Assessing the added value of wastewater-based epidemiology to monitor illicit drug use. Thesis, University of Lausanne Open Archive. Retrieved from http://serval.unil.ch Document URN: urn:nbn:ch:serval-BIB_F480FD9ACCB05.

Bishop, N., Jones-Lepp, T., Margetts, M., Sykes, J., Alvarez, D., & Keil, D. E. (2020). Wastewater-Based Epidemiology Pilot Study to Compare Drug Use in Rural and Urban Areas in the Western United States (manuscript is in submission to Sci Total Environ)

Bruno, R., Edirisinghe, M., Hall, W., Mueller, J., Lai, F., O’Brien, J., & Thai, P. (2018). Association between purity of drug seizures and illicit drug loads measured in wastewater in a South East Queensland catchment over a six year period. Science of the Total Environment, 635, 779–783.

Castiglioni, S., Bijlsma, L., Covaci, A., Emke, E., Hernández. F., Reid, M., Ort, C., Thomas, K. V, van Nuijs, A. L, de Voogt, P., Zuccato, E. (2013). Evaluation of uncertainties associated with the determination of community drug use through the measurement of sewage drug biomarkers. Environmental Science & Technology, 47(3), 1452–1460.

Castiglioni, S., Thomas, K.V., Kasprzyk-Hordern, B., Vandam, L., & Griffiths, P. (2014). Testing wastewater to detect illicit drugs: State of the art, potential and research needs. Science of the Total Environment, 487, 613–620.

Castiglioni, S., & Vandam, L. (2016). “A global overview of wastewater-based epidemiology”, pp. 45–54 in Assessing illicit drugs in wastewater: Advances in wastewater-based drug epidemiology, EMCDDA Insights 22, Publications Office of the European Union, Luxembourg.

Centers for Disease Control and Prevention (CDC). (2017). What States Need to Know about PDMPs. Retrieved from https://www.cdc.gov/drugoverdose/pdmp/states.html

Chen, C., Kostakis, C., Gerber, J., Tscharke, B., Irvine, R., & White, J. (2014). Towards finding a population biomarker for wastewater epidemiology studies. Science of the Total Environment, 487(1), 621–628.

Daughton, C. (2001). Emerging pollutants, and communicating the science of environmental chemistry and mass spectrometry: Pharmaceuticals in the environment. Journal of the American Society for Mass Spectrometry, 12(10), 1067–1076.

Davenport, S., Weaver, A., Caverly, M. (Society of Actuaries). (2019). Economic Impact of Non-Medical Opioid Use in the United States Annual Estimates and Projections for 2015 through 2019. Retrieved from https://www.soa.org/globalassets/assets/files/resources/research-report/2019/econ-impact-non-medical-opioid-use.pdf

Department of Homeland Security. (2015). National Infrastructure Protection Plan (NIPP) Water and Wastewater Sector-Specific Plan for 2015. Retrieved from https://www.cisa.gov/publication/nipp-ssp-water-2015

Dunn, K. E., Barrett, F. S., Yepez-Laubach, C., Meyer, C., Hruska, B., Petrush, K., … Bigelow, G. (2016). Opioid Overdose Experience, Risk Behaviors, and Knowledge in Drug Users from a Rural versus an Urban Setting. Journal of Substance Abuse Treatment, 71, 1–7. doi: 10.1016/j.jsat.2016.08.006.

Hall, W., Prichard, J., Kirkbride, P., Bruno, R., Thai, P. K., Gartner, C., … Mueller, J. F. (2012). An analysis of ethical issues in using wastewater analysis to monitor illicit drug use. Addiction, 107, 1767–1773.

Jeffery, M. M., Hooten, W. M., & Henk, H. J. Bellolio, M., Hess, E.P., Meara, E., … Shah, D. (2018). Trends in opioid use in commercially insured and Medicare Advantage populations in 2007–16: Retrospective cohort study. BMJ 2018;362:k2833. doi:10.1136/bmj.k2833 pmid:30068513.

Keshaviah, Aparna (ed.). The Potential of Wastewater Testing for Public Health and Safety.Washington, DC: Mathematica Policy Research, August, 2017. Retrieved from https://www.mathematica.org/our-publications-and-findings/publications/the-potential-of-wastewater-testing-for-public-health-and-safety-special-report Accessed February 19, 2020.

McCall, A., Bade, R., Kinyua, J., Yin Lai., F., Thai, P., Covaci, A., … Ort, C. (2016). Critical review on the stability of illicit drugs in sewers and wastewater samples. Water Research, 88, 933–947.

Montana Department of Justice. (2017). Montana Department of Justice Forensic Science Division Annual Report–2017. Retrieved from https://media.dojmt.gov/wp-content/uploads/2017-FSD-Annual-Report-2.pdf

Montana Medical Association, Department of Public Health and Human Services. (2015). Opioid use statistics. Retrieved from http://knowyourdosemt.org/wp-content/uploads/2015/03/Neonatal-Abstinence-Syndrome-in-Montana-Newborns-2000-2013.pdf

National Institute of Drug Abuse. (2018). Montana Opioid Summary: Opioid Related Overdose Deaths. Retrieved from https://www.drugabuse.gov/drugs-abuse/opioids/opioid-summaries-by-state/montana-opioid-summary

Thiels, C.A., Habermann, E. B., Hooten, W. M., & Jeffery, M. M. (2019). Chronic use of tramadol after acute pain episode: Cohort study. British Medical Journal. 365, L1849.

Van Nuijs, A., Covaci, A., Beyers, H., Bervoets, L., Blust, R., Verpooten, G., … Jorens, P. (2015). Do concentrations of pharmaceuticals in sewage reflect prescription figures? Environmental Science and Pollution Research International, 22(12), 9110–9118.

Van Nuijs, A., Mougel, J., Tarcomnicu, I., Bervoets, L., Blust, R., Jorens, P., … Covaci, A. (2011). Sewage epidemiology: A real-time approach to estimate the consumption of illicit drugs in Brussels, Belgium. Environment International, 37(3), 612–621.

Zuccato, E., Chiabrando, C., Castiglioni, S., Bagnati, R., & Fanelli, R. (2008). Estimating community drug abuse by wastewater analysis. Environmental Health Perspectives, 116(8), 1027–1032.

